# Detecting Basilar Artery Wall Pathology in Isolated Pontine Infarction with Short-TE SWI Magnitude Images

**DOI:** 10.1101/2025.09.23.25336518

**Authors:** Hon-Man Liu, Adam Huang, Yung-Chuan Huang, Chih-Hui Wu, Li-Shan Hsu, Ning Yang, Kuo-Chi Lin

## Abstract

**Background and Purpose:** Intracranial atherosclerosis is a significant contributor to ischemic stroke, often through plaques without luminal stenosis. Current imaging techniques struggle to detect vessel wall pathology effectively. This study evaluates short-TE susceptibility-weighted imaging magnitude images (STE-MI) for identifying cerebral microbleeds and basilar artery (BA) wall changes in acute isolated pontine infarction, with reduced artifacts compared to SWAN.

**Methods:** In this post hoc analysis, 2,475 brain MR exams conducted between March 2022 and March 2025 using a 3.0 T scanner compared STE-MI (TE=7.2 ms) with SWAN for microbleed detection and BA artifact reduction. Among 131 patients with acute posterior circulation stroke (January 2023–April 2025), vessel wall pathology was assessed in 92 patients with vertebrobasilar stenosis (VBASO) and 39 with non-stenotic non-cardioembolic (NSNCE) stroke using STE-MI. Interobserver agreement and comparative statistics were analyzed.

**Results:** STE-MI detected 86% of microbleeds identified by SWAN in 33 subjects (Cohen’s kappa=0.828 for interobserver agreement). Blooming artifacts around the BA were reduced in 96.9% of 2,475 cases with STE-MI compared to 5.5% with SWAN. In the NSNCE group, 10 of 16 patients with isolated pontine infarction exhibited extraluminal BA pathology on STE-MI (mixed hypo-/intermediate intensity in 6, hypointensity in 4), correlating with diffusion-weighted imaging (DWI) lesions.

**Conclusions:** STE-MI effectively minimizes artifacts and detects BA wall pathology in acute isolated pontine infarction, particularly in NSNCE stroke. This approach may enhance the identification of culprit lesions, potentially improving risk stratification and guiding targeted therapies.

## Introduction

Intracranial atherosclerosis is a critical factor in ischemic stroke, characterized by plaque accumulation within intracranial arteries, leading to vessel narrowing and reduced cerebral blood flow. The TOAST classification categorizes ischemic stroke into five subtypes based on etiology: large artery atherosclerotic stenosis, small artery disease, major-risk source cardiogenic embolism, cryptogenic stroke, and unusual causes (e.g., arterial dissection) [1]. A systematic review [2] reported that culprit intracranial plaques without substantial stenosis were present in 51.2% of acute ischemic strokes detected by vessel wall MRI. Many researchers argue that current vascular assessments primarily focus on the vessel lumen, while atherosclerotic plaques originate in the vessel wall and may cause ischemic stroke even without luminal stenosis [3,4].

The clinical concept of embolic stroke of undetermined source (ESUS) [5], accounting for 20% to 30% of all ischemic strokes, aims to identify patients with non-lacunar strokes without proximal arterial stenosis or a major cardiac embolic source. There is a complex relationship between calcification, intraplaque hemorrhage (IPH), and lipid core within atherosclerotic plaques, which are associated with plaque vulnerability [6]. Calcified plaques tend to be more stable and less prone to rupture compared to non-calcified plaques. The presence of IPH indicates an unstable plaque, as bleeding into the plaque often precedes rupture. High-resolution vessel wall MRI (HRVW-MRI) sequences help differentiate tissue types based on their relaxation times. Calcification typically appears hypointense on both T1-and T2-weighted images. Lipid-rich areas and subacute IPH usually appear hyperintense on T1-weighted images, while fibrous tissue may present differently on T2-weighted images [7,8].

Susceptibility-weighted imaging (SWI) is a specialized sequence more sensitive than conventional MRI in detecting brain calcium and iron, which distort the local magnetic field [9]. Compared with CT, SWI is not inferior in detecting intracranial vertebral artery (VA) calcification [10] and may be superior in identifying non-stenotic atherosclerosis, mural hematoma, or dissection [10,11]. SWAN, a specific pulse sequence using a multi-TE readout technique, generates a higher signal-to-noise ratio and is less affected by chemical shift compared to conventional SWI techniques. However, SWAN is limited by blooming artifacts around the BA, hindering accurate assessment of vessel wall pathology in non-stenotic stroke [11]. Magnitude images (MI) in SWI are generated based on the signal intensity of different tissues, reflecting their magnetic susceptibility properties. Areas with high susceptibility appear darker due to local magnetic field distortions. Several factors influence MI in SWI, including T2* relaxation time of tissues, magnetic susceptibility differences, magnetic field strength, acquisition parameters, phase cancellation, and post-processing techniques [12]. Echo time (TE) is crucial for optimizing image quality, contrast, and sensitivity to various tissues and pathologies in SWI. Shorter TEs (STE) may provide higher signal intensity but might not fully capture susceptibility effects. For detecting acute hemorrhages, a shorter TE may be preferred to capture the initial signal before significant T2* decay reduces intensity. Conversely, longer TEs can enhance sensitivity to susceptibility effects, improving visualization of small veins, microbleeds, and iron deposits.

We hypothesized that STE-MI, with reduced TE, minimizes artifacts and enhances detection of BA wall pathology in acute isolated pontine infarction, particularly in non-stenotic non-cardioembolic (NSNCE) stroke. The objectives of this study were: To compare the feasibility and capability of detecting cerebral microbleeds or calcification between the MI of short-TE-SWI (STE-MI) and conventional SWAN, to demonstrate that STE-MI can reduce blooming artifacts on the BA from adjacent bones and air, as well as flow artifacts seen on SWAN. to explore the application of STE-MI in patients with acute isolated pontine infarction.

## Materials and Methods

### Ethics Statement

The study was approved by the local ethics committee and conducted in accordance with the principles of the Declaration of Helsinki.

### Study Design and Imaging Methods

This post hoc analysis included brain MR examinations performed between March 21, 2022, and March 20, 2025, using a 3.0 T scanner (Ingenia, Philips). The protocol comprised DWI, T2-FLAIR, 3D-SWI (SWAN), black-blood 1 mm T1-weighted imaging, TOF or contrast-enhanced MRA, and post-contrast enhanced black-blood 1 mm T1-weighted imaging. SWAN parameters were: TR = 31 ms; TE = 7.2, 13, 20, and 26 ms; flip angle = 17°; matrix = 384×384; FOV = 230×230 mm; slice thickness = 2 mm; interslice spacing = 1 mm; and compressed sense factor = 2. The parameters for STE-SWI were identical to SWAN, except for using a single short TE. In SWAN, the imaging output included four sets of the MI with different TEs, four unwrapped phase images (PI), and one combination-filtered phase image. The STE-MI consisted of the first two sets of MI with short TE in SWAN, using TEs of 7.2 and 13 ms, respectively, based on our pilot study. Additionally, black-blood 1 mm T1-weighted images (BRAINVIEW on our machine) were routinely obtained with parameters: TR = 600 ms; TE = 30 ms; matrix: 270×270 mm; FOV = 208×208 mm; flip angle = 90 degrees; slice thickness = 1 mm; and spacing = 1 mm.

Exclusion criteria included poor image quality, incomplete data, patients under 20 years of age, and obvious artifacts on the BA wall from other sources, such as compressed sense or flow artifacts.

### Study Components

#### Cerebral Microbleed Detection Comparison

Microbleeds were defined as signal void areas in the brain parenchyma with a diameter of 2-10 mm and blooming effects [13, 14]. Using 33 subjects, two board-certified radiologists independently compared the detection capabilities of SWAN and STE-MI. They evaluated conspicuity using a five-grade scale ranging from “SWAN much better” to “much worse” than STE-MI.

#### Artifact Reduction Assessment

Between March 21, 2022, and March 20, 2025, 2,475 subjects were evaluated to compare BA visualization between SWAN and STE-MI. Visualization quality was classified as: Excellent: 360-degree BA wall visualization; Good: >180 but <360-degree visualization; Poor: <180-degree visualization

#### Clinical Application of STE-MI in Patients with Isolated Pontine Infarction

From January 1, 2023, to April 30, 2025, 131 patients with acute posterior circulation ischemic stroke were identified by diffusion-weighted imaging (DWI) using a 3.0 T MR scanner at our institute. Vessel wall changes were analyzed with STE-MI in 92 patients with stenotic vertebrobasilar artery (VBA) and 39 patients with NSNCE stroke on MRA. BA stenosis was assessed on 3-dimensional time-of-flight MRA, contrast-enhanced MRA using source images and 3-dimensional reconstructed views, and the axial section of the BA lumen for luminal stenosis. Significant stenosis in the VBASO group was defined as luminal narrowing greater than 30% [15]. All patients in the NSNCE group had isolated pontine infarction without significant BA stenosis on MRA and no evidence of atrial fibrillation on 24-hour Holter monitoring. However, other possible embolic sources, such as patent foramen ovale or aortic diseases, could not be completely excluded.

Isolated pontine infarction was defined as ischemic stroke characterized by a small, well-circumscribed area of infarct in the pons without other posterior circulation stroke. Based on the territory of the pontine artery, unilateral pontine infarction was classified into five subtypes: lateral, anterolateral, paramedian, tegmental, and small deep pontine infarct, according to the lesion location on DWI [16–18]. Tegmental and small deep pontine infarcts were defined as unilateral pontine infarcts not involving the ventral surface of the pons, while others involving the ventral surface were named according to their location. Lesion distribution was evaluated by dividing the cross-section of the BA lumen into four equal arcs (right lateral, ventral, left lateral, and dorsal) on axial SWI at the level or just proximal to the lesion shown on DWI [19]. Demographic and clinical information was recorded on admission, including age, gender, stroke risk factors (hypertension, diabetes mellitus, hyperlipidemia, smoking, and atrial fibrillation), and history of medication (anti-hypertensives, antiplatelets, and statins). Hypertension was defined as a history of antihypertensive medication use or systolic blood pressure (BP) >140 mmHg or diastolic BP >90 mmHg at discharge. Systolic and diastolic blood pressure were measured from the arm in a supine position on admission. Diabetes was defined as the use of hypoglycemic agents, fasting blood glucose >126 mg/dl, random blood glucose >200 mg/dl, or glycosylated hemoglobin >6.4% on admission. Hyperlipidemia was defined as prior use of lipid-lowering agents, fasting serum total cholesterol level >240 mg/dl, and/or low-density lipoprotein level >160 mg/dl. Laboratory tests were conducted at admission. Lipid profiles, including total cholesterol, high-density lipoprotein, low-density lipoprotein, triglycerides, fasting blood glucose, and hemoglobin A1C, were measured the day after admission following overnight fasting.

Subjects were classified as current smokers or non-smokers. To investigate atrial fibrillation, patients underwent 24-hour Holter monitoring. Risk factors were also examined using the machine learning model XGBoost [20] (with default parameters).

### Statistical Analysis

Cohen’s kappa was used to assess interobserver agreement for microbleed detection. Agreement levels were categorized as: <0.20: poor; 0.21-0.40: fair; 0.41-0.60: moderate; 0.61-0.80: substantial; 0.81-1.00: almost perfect. Differences between variables were tested using Pearson’s chi-square test, Fisher’s exact test, Student’s t-test, or the Mann-Whitney U test, as appropriate. Data were analyzed using R software, version 3.5.1 for Windows. A p-value less than 0.05 was considered statistically significant.

Using the machine learning model XGBoost, feature importance was evaluated using the “gain” metric, where a higher score indicates a greater impact on model performance. XGBoost was selected for its effectiveness in exploring complex data relationships, its use of regularization techniques to prevent overfitting, and its robust handling of missing values.

## Results

### Study Population

A total of 2,662 examinations using a 3.0 T MR scanner were conducted during the study period. Of these, 187 studies were excluded based on the exclusion criteria. The indications for the remaining 2,475 examinations included cerebrovascular diseases (n=726), tumor staging and post-treatment follow-up (n=652), headache (n=350), dementia (n=276), dizziness (n=184), seizure (n=45), suspected intracranial infectious/inflammatory disease (n=55), and other indications (n=87).

### Cerebral Microbleed Detection Comparison

In 33 cases with cerebral microbleeds, 30 slices were analyzed. SWAN identified 160 microbleeds per formal MR reports. Two readers detected 138 (86.3%) and 137 (85.6%) microbleeds on STE-MI, respectively, with excellent interobserver agreement (Cohen’s kappa=0.828, 95% CI: 0.600-1.000). STE-MI was more likely to miss smaller hypointense lesions (Supplementary Fig S1).

### Artifact Reduction Assessment

Blooming artifacts from air and bone around the posterior fossa, particularly the basilar artery (BA), increased with longer TE on SWAN (Fig 1). In 2,475 exams, STE-MI reduced artifacts and provided excellent BA visualization in 96.9% of cases, compared to 5.5% with SWAN (p < .0001; Table 1). Optimal TE for balancing image quality and hypointensity detection was 7.2-13 ms. TE values over 20 ms yielded minimal good-quality BA imaging.

**FIG. 1:**
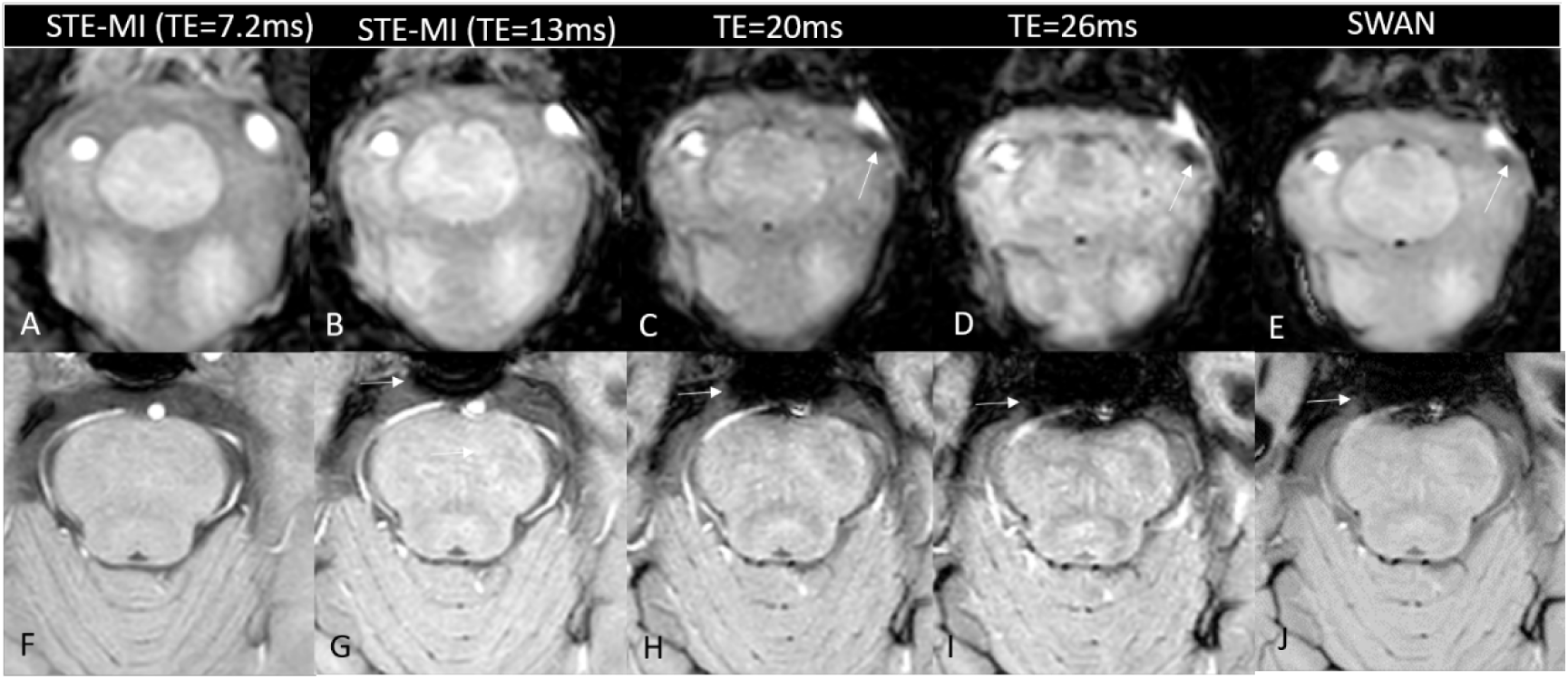
STE-MI at TE=7.2 ms optimizes basilar artery (BA) wall visualization for pathology detection. This figure compares blooming and phase artifacts at varying echo times (TE) at the foramen magnum (upper row) and mid-pons (lower row) levels. Bone-air artifacts increase significantly when TE exceeds 13 ms. Phase artifacts near vessel segments (white arrow; panels C, D, E, upper row) intensify with longer TE in the left vertebral artery (VA) due to its oblique orientation. STE-MI (panels A and F; TE=7.2 ms) best differentiates vessels (VA and BA), cerebrospinal fluid (CSF), and brainstem structures.

**Table 1.**
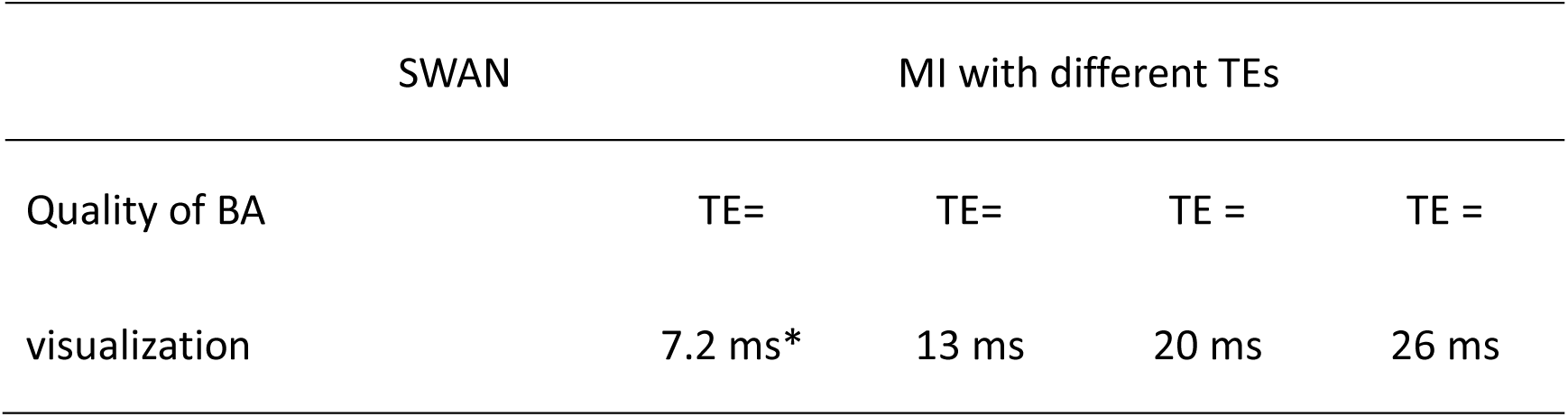
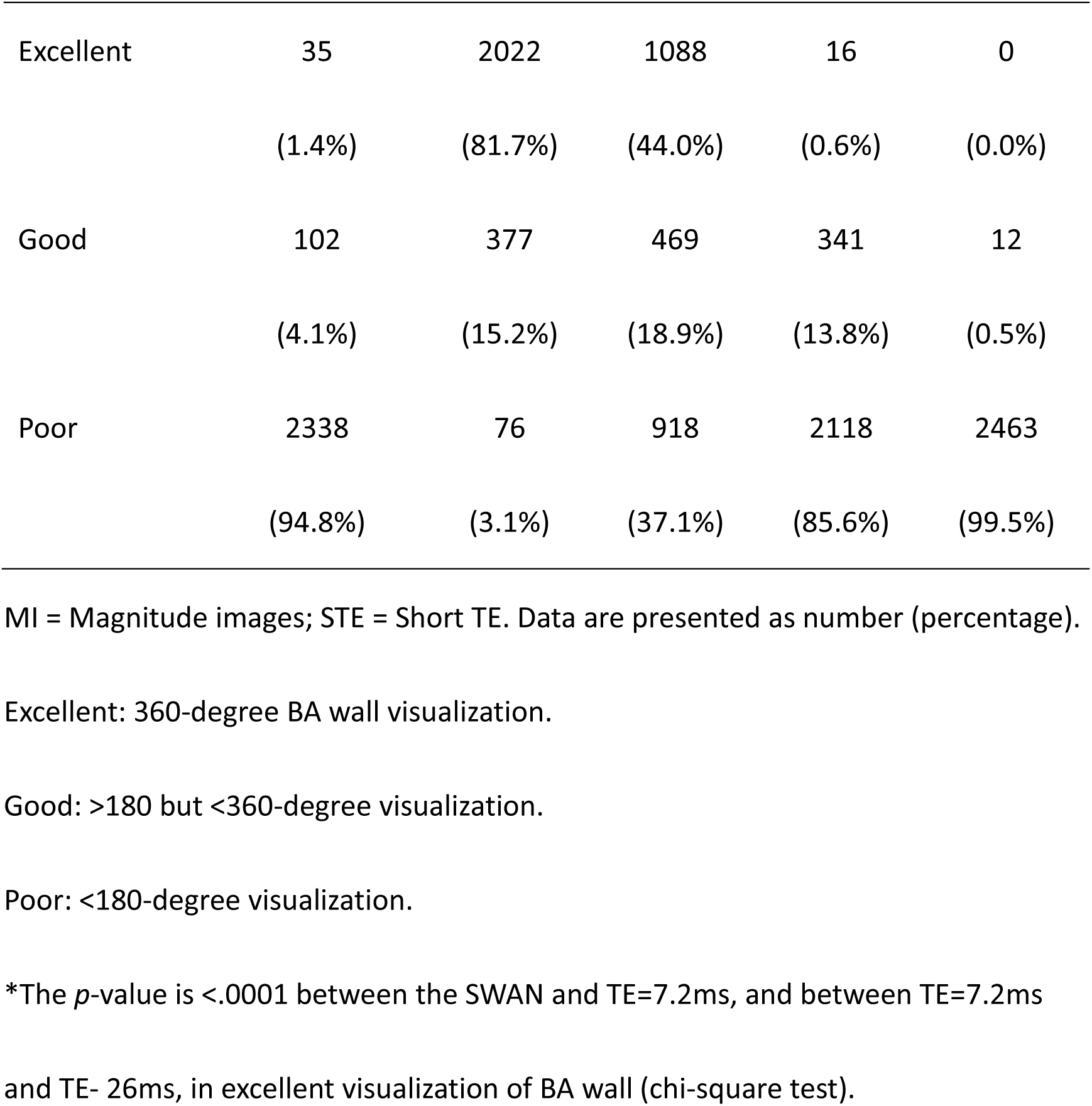
Comparison of blooming artifacts obscuring the BA from adjacent bone and air between from SWAN and magnitude images with different TEs in 2,475 examinations.

### Clinical Application of STE-MI in Patients with Isolated Pontine Infarction

Of 131 patients with acute posterior circulation stroke from January 2023 to April 2025, 92 had vertebrobasilar artery stenosis/occlusion (VBASO), with 13 showing isolated pontine lesions on DWI. Of 39 NSNCE stroke patients, 16 had isolated pontine lesions. Clinical characteristics differed significantly in current smoking (p=0.041) and low serum triglycerides (p=0.033) in the NSNCE group (Supplementary Table S1). XGBoost analysis highlighted triglycerides and smoking as key risk factors for NSNCE pontine infarction (Supplementary Fig S2).

On STE-MI (Fig 1), BA lumen appeared hyperintense without wall thickening; abnormal iso-/hypointensity was conspicuous outside the lumen. In occlusion or stenosis, luminal hyperintensity was reduced or absent. Among 16 NSNCE pontine infarct patients, lesions were on the right side in 5, with SWI-MI findings on dorsal (n=7), left lateral (n=5), right lateral (n=2), and ventral (n=1) BA aspects. Abnormal BA wall signals were seen in 10 (62.5%) patients on STE-MI: mixed hypo-/intermediate intensity in 6 (60.0%) and hypointensity in 4 (40.0%) (Fig 2). These extraluminal outward-protruding changes, presumed culprit lesions, matched DWI lesion locations. One patient showed hypointensity in the posterior cerebral artery wall without CT calcification (Supplementary Fig S3). SWAN phase images, when available, distinguished calcification (high signal) from hemorrhage (low signal) (Figs 3, 4, 5, 6). Black-blood 1 mm T1-weighted imaging showed thickened walls in 4 of 16 patients, with no abnormal enhancement.

**FIG. 2:**
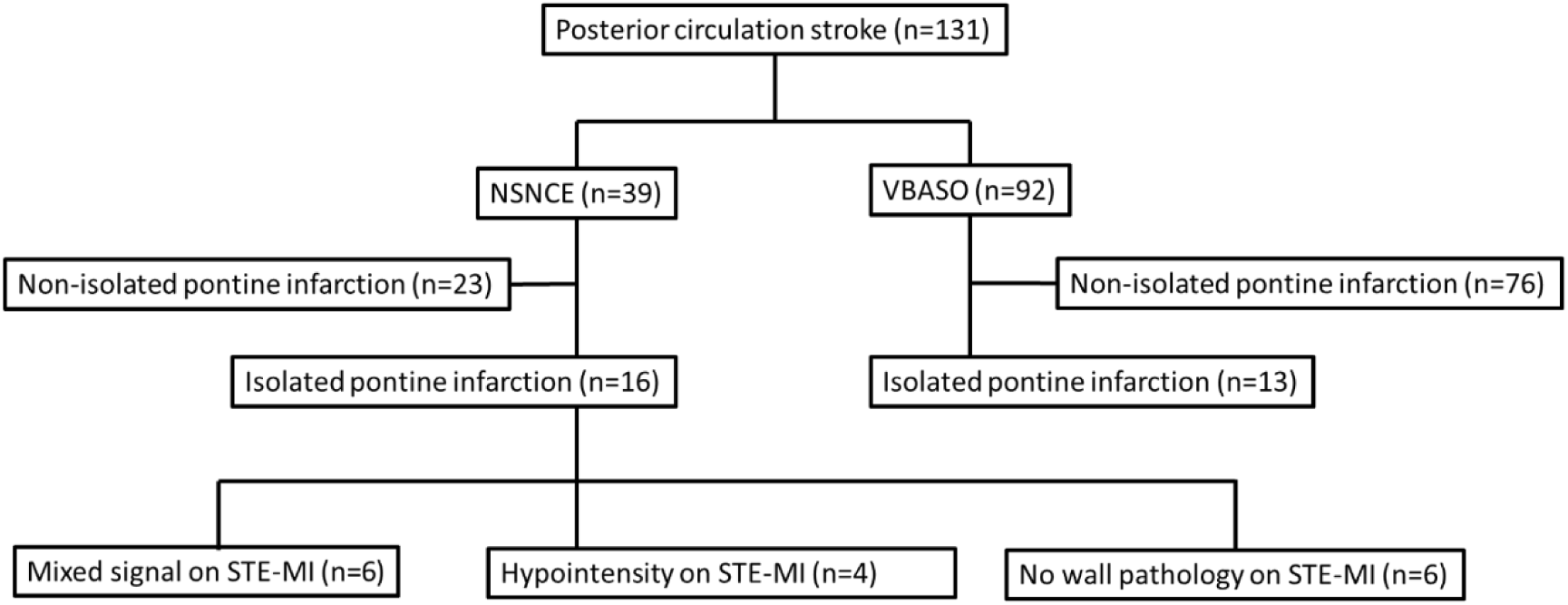
Distribution of 131 patients with posterior circulation stroke: 92 with vertebrobasilar artery stenosis or occlusion (VBASO) and 39 with non-stenotic non-cardioembolic (NSNCE) stroke. Isolated pontine infarcts are noted in 13 VBASO and 16 NSNCE patients. On STE-MI, 6 NSNCE patients show no wall pathology, 6 have mixed signal intensity pathology, and 4 exhibit hypointensity.

**FIG. 3:**
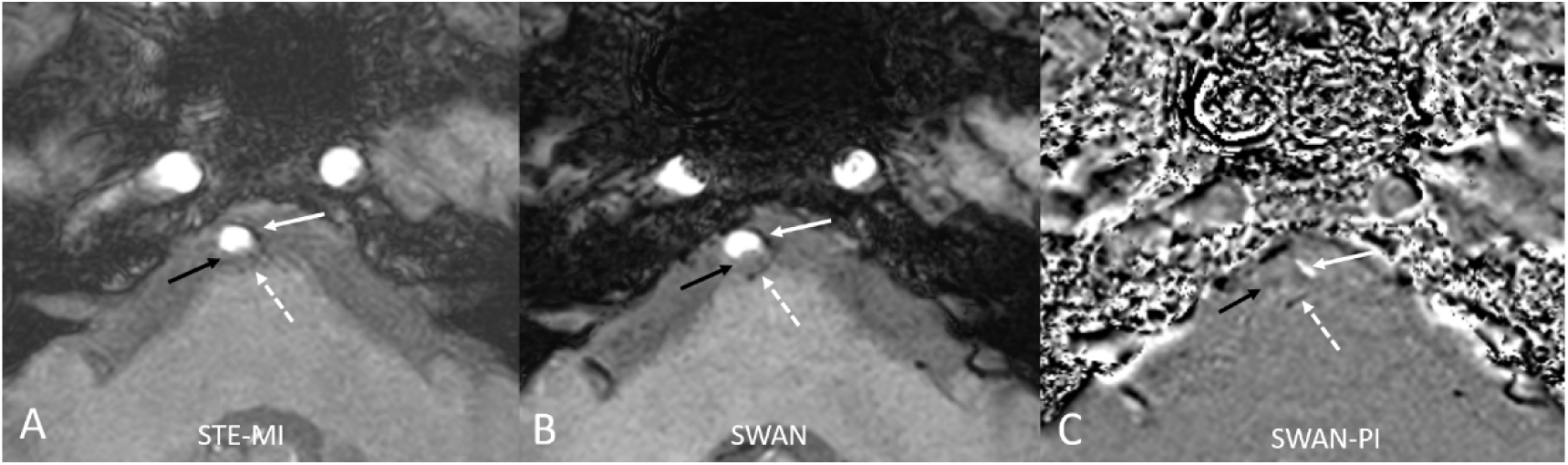
Comparative analysis of TE effects on STE-MI (panel A), SWAN (panel B), and corresponding phase image (PI) (panel C). Minimal artifacts from clivus and bone allow clear BA visualization on both. An intermediate signal (black arrow) appears on the dorsal BA wall in both STE-MI and SWAN. A hypointense rim (dashed white arrow) is more conspicuous on SWAN, suggesting hemorrhage. Hypointensity on the left BA wall (white arrow) appears hyperintense on SWAN-PI (panel C), indicating calcification.

Pontine DWI lesion distribution in 16 NSNCE patients included right lateral (n=1), right anterolateral (n=2), right paramedian (n=5), left paramedian (n=8), left anterolateral (n=2), tegmental (n=2), and small deep pontine infarct (SDPI, n=2), totaling 22 lesions (6 patients had multiple lesions). On STE-MI, extraluminal pathology appeared in 15 BA arcs: right lateral (n=2), ventral (n=1), left lateral (n=6), and dorsal (n=8). Six patients had no obvious wall lesions; five had two arcs involved (Fig 7).

In 13 VBASO pontine infarct patients, 9 lesions were on the right side, with SWI-MI findings on dorsal (n=6), left lateral (n=2), right lateral (n=4), and ventral (n=2) BA aspects. Mixed hypo-/intermediate intensity, likely from blood clots and atheromas, appeared in 11 patients at steno-occlusion sites. Circular voids or intermediate changes matched DWI lesions in 2 patients. Black-blood 1 mm T1-weighted imaging showed thickened walls in 4 patients, with contrast enhancement in 1.

## Discussion

In this study, we found that approximately 80–96% of microbleeds larger than 2 mm on SWAN were also conspicuous on STE-MI, with excellent interobserver agreement (Cohen’s kappa = 0.828). Blooming artifacts affecting the BA were eliminated in 96.9% of cases using STE-MI (TE = 7.2 ms), compared to only 5.5% with SWAN, providing superior visualization of the BA wall. Among 131 patients with acute posterior circulation ischemic stroke, 16 NSNCE stroke patients had isolated pontine infarction, and 10 (62.5%) exhibited BA extraluminal pathology on STE-MI (mixed hypo-/intermediate intensity in 6, hypointensity in 4). These pathologies corresponded to DWI lesion locations and were presumed to be culprit lesions. Black-blood 1 mm T1-weighted imaging was less effective in detecting BA wall pathology, a finding rarely mentioned in the literature that may partly explain NSNCE etiology.

Literature indicates that IPH often appears hyperintense on T1-weighted HRVW-MRI due to methemoglobin [20], but this is limited to the subacute stage. HRVW-MRI has drawbacks, including longer scan times and insensitivity to calcification [11]. SWI, particularly STE-MI, excels in detecting cerebral hemorrhage and calcification, identifying lesions ≥2 mm. STE-MI reduced artifacts from adjacent structures, enabling evaluation of BA wall pathology in isolated pontine infarction. Over half of ischemic stroke patients lack significant stenosis or clear etiology, with low-degree BA stenosis more prevalent than in the MCA [2], and IPH prevalence similar across stenosis grades [22]. Recent studies on posterior circulation plaques highlight features like geometry and intraplaque hemorrhage [23, 24]. Our focus on extraluminal outward-protruding pathology is novel; such plaques may occlude perforators despite small size, as confirmed by their correlation with DWI lesions.

STE-MI identified microcalcification (Figs 3, 4), IPH (Figs 4, 5), and non-calcified components (Figs 5, 6) as culprit lesions when phase images were available, crucial for risk stratification and therapy guidance.

**FIG. 4:**
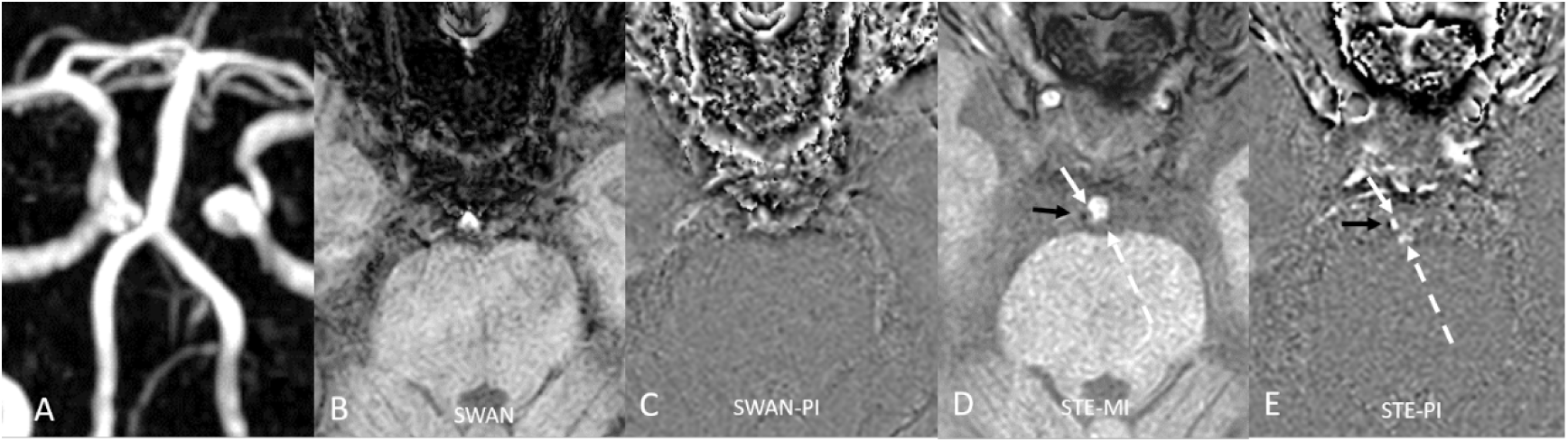
Comparison of SWAN and STE-SWI in an NSNCE BA stroke patient. Panel A shows normal BA on MR angiography. Panels B-E display SWAN (B), SWAN-PI (C), STE-MI (D), and STE-PI (E). SWAN artifacts obscure BA, while STE-MI offers clear visualization. STE-MI (panel D) reveals a hypointense spot (black arrow) and mild intermediate intensities (white and dashed arrows) on the right and posterior BA walls. On STE-PI (panel E), hypointensity suggests hemorrhage; intermediate intensities indicate calcification, marking potential plaque instability.

**FIG. 5:**
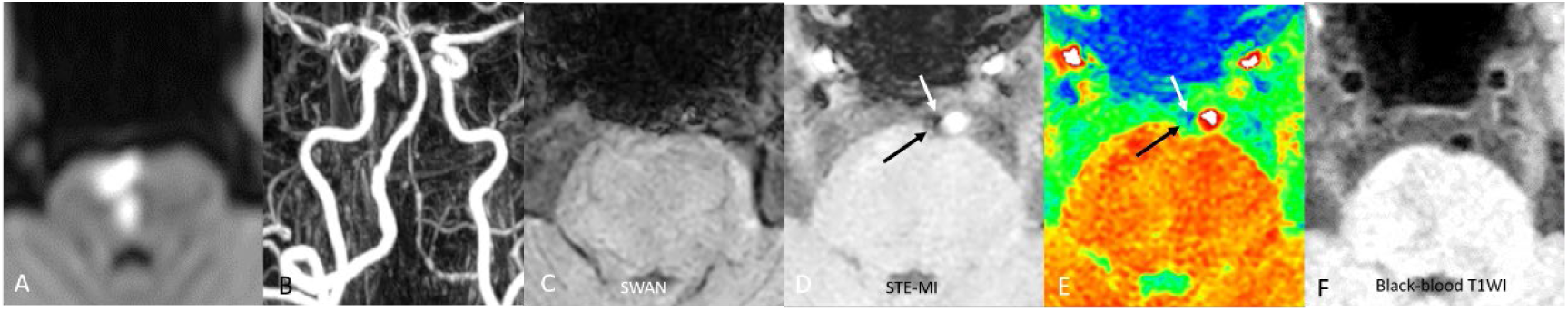
Imaging findings in an acute NSNCE BA stroke patient. Diffusion-weighted imaging (DWI, panel A) shows acute ischemic stroke in the right paramedian pons. MR angiography (panel B) indicates no significant BA stenosis. SWAN (panel C) artifacts obscure BA, while STE-MI (panel D) reveals focal hypointensity (white arrow) and intermediate signal (black arrow) on the right BA wall, delineated on color mapping (panel E) as possible hemorrhage or calcification. Black-blood 1 mm T1-weighted images (panel F) show no abnormal wall signal or enhancement.

**FIG. 6:**
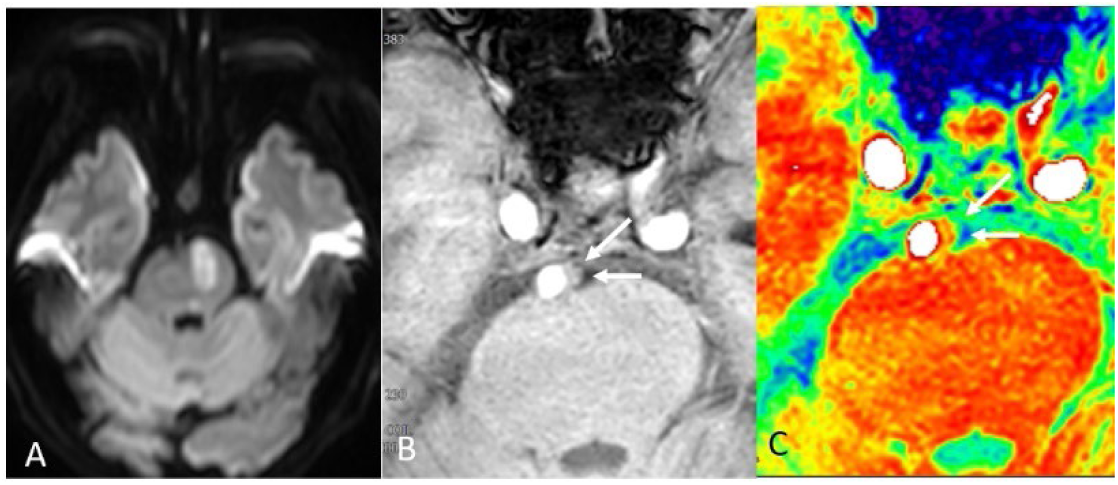
Imaging of a male patient with acute NSNCE left pontine ischemia. DWI (panel A) shows hyperintensity in the left paramedian pons. STE-MI (panel B) reveals hypointensity (arrows) on the left BA wall with an adjacent isointense area. Color mapping (panel C) suggests a plaque with calcified, non-calcified, and hemorrhagic components, indicating potential plaque instability.

**FIG. 7:**
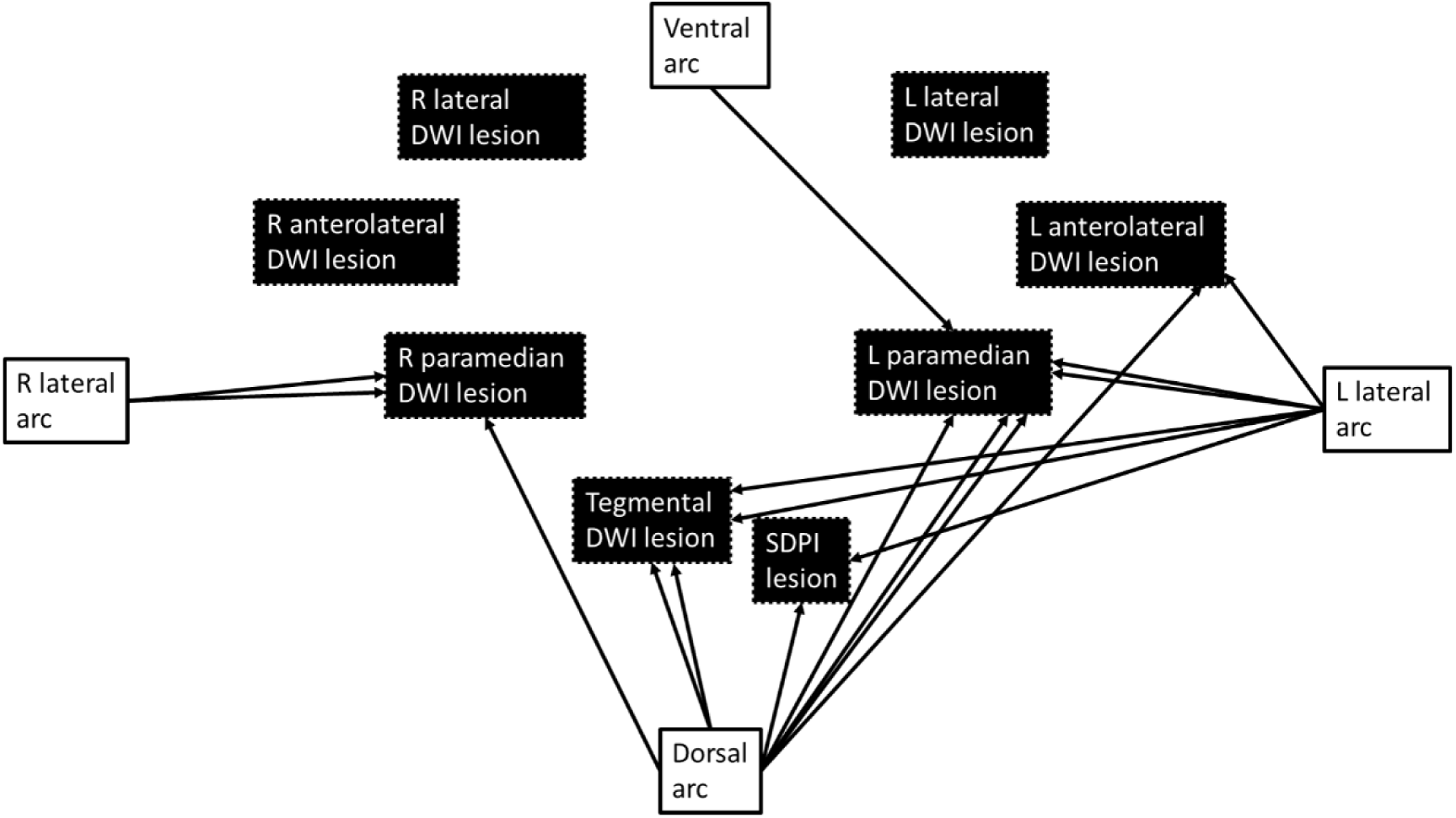
Relationship between DWI lesion locations and extraluminal outward-protruding pathology distribution in the BA lumen cross-section in 13 NSNCE isolated pontine infarction patients. Arrows mark pathology locations, potentially culprit lesions for corresponding DWI lesions. Six patients show no obvious pathology; five have two arcs involved. R: right, L: left; DWI: diffusion-weighted imaging, SDPI: small deep pontine infarct.

Autopsy reports suggest this extraluminal pathology is unlikely related to VBA outward remodeling [25] but may reflect atherosclerosis progression in the tunica adventitia, which actively contributes to plaque development [25, 26]. Adventitial thickening correlates with intimal changes [27], and vasa vasorum facilitate neovascularization in plaques [28]. Imaging the adventitia offers new insights into atherosclerosis risk [29], and STE-MI may serve as a non-invasive tool for assessing extraluminal pathology.

We used “non-stenotic non-cardioembolic (NSNCE)” instead of ESUS, though half our patients were diagnosed as ESUS. While screened for atrial fibrillation, many lacked full evaluation for conditions like patent foramen ovale. NSNCE patients showed extraluminal pathology, often hypointense on SWI, suggesting calcification or hemorrhage akin to atherosclerotic plaques. This pathology, likely outward-protruding plaques, differs from inward-protruding stenotic plaques and may cause emboli or perforator occlusion.

ESUS management focuses on secondary prevention, with routine anticoagulation not universally beneficial [30, 31]. Subgroups like those with atrial cardiopathy may benefit from anticoagulation (Apixaban) over antiplatelet therapy (Aspirin) [32, 33]. Our findings suggest another subgroup with outward-protruding plaques may benefit from antiplatelets or statins. Subclassifying NSNCE patients with such pathology could enhance risk stratification and guide personalized therapies, potentially impacting clinical practice if antiplatelet optimization proves superior to anticoagulation.

Limitations include lack of histological correlation, small sample size, and insufficient resolution for normal vessel wall thickness. This study focused solely on the BA, excluding the MCA. Comparison with HRVW-MRI in larger series is ongoing. Phase image generation was challenging for STE-SWI and SWAN, and mixed calcification/hemorrhage in vessel walls suggests quantitative susceptibility mapping as a potential solution. A larger prospective study with treatments like statins for this group is warranted.

## Conclusion

STE-MI can detect magnetic susceptibility changes in the extraluminal outward-protruding pathology on the BA that were not identified by other MR studies. Such changes may theoretically be due to either hemorrhagic products or calcification. The clinical significance of magnetic susceptibility changes in NSNCE stroke warrants further investigation.

## Data Availability

The data that support the findings of this study are available from the corresponding author, HML, upon reasonable request.

## Non-standard Abbreviations and Acronyms

BA: Basilar artery
DWI: Diffusion-weighted imaging
ESUS: Embolic stroke of undetermined source
HRVWI-MRI: High-resolution vessel wall
MRI IPH: Intraplaque hemorrhage
MI: Magnitude images
NSNCE: Non-stenotic non-cardioembolic
PI: Phase images
STE: Shorter TEs
STE: MI: short-TE susceptibility-weighted imaging magnitude images
SWI: Susceptibility-weighted imaging
TE: Echo time
VA: Vertebral artery
VBA: Vertebrobasilar artery
VBASO: Vertebrobasilar artery stenosis/occlusion

## Acknowledgements

Grant support: This work is supported by grants from the Fu Jen Catholic University Hospital (PL-202308050-V to Hon-Man Liu).

